# Symptom profiles and accuracy of clinical definitions for COVID-19 in the community. Results of the Virus Watch community cohort

**DOI:** 10.1101/2021.05.14.21257229

**Authors:** Ellen Fragaszy, Madhumita Shrotri, Cyril Geismar, Anna Aryee, Sarah Beale, Isobel Braithwaite, Thomas Byrne, Wing Lam Erica Fong, Jo Gibbs, Pia Hardelid, Jana Kovar, Vasileios Lampos, Eleni Nastouli, Annalan M D Navaratnam, Vincent Nguyen, Parth Patel, Robert W Aldridge, Andrew Hayward, on behalf of Virus Watch Collaborative

## Abstract

**Background:** Understanding the symptomatology and accuracy of clinical case definitions for COVID-19 in the community is important for the initiation of Test, Trace and Isolate (TTI) and may, in future, be important for early prescription of antivirals.

**Methods:** Virus Watch is a large community cohort with prospective daily recording of a wide range of symptoms and self-reporting of swab results (mainly undertaken through the UK TTI system). We compared frequency, severity, timing, and duration of symptoms in test positive and test negative cases. We compared the test performance of the current UK case definition used by TTI (any one of: new continuous cough, high temperature, or loss of or altered sense of smell or taste) with a wider definition that also included muscle aches, chills, headache, or loss of appetite.

**Findings:** We included results from 8213 swabbed illnesses, 944 of which tested positive for SARS-CoV-2. All symptoms were more common in test positive than test negative illnesses and symptoms were also more severe and of longer duration. Common symptoms such as cough, headache, fatigue, muscle aches, and loss of appetite occurred early in the course of illness but were also very common in test-negative illnesses. In contrast, high temperature and loss of or altered sense of smell or taste were less frequently identified in swab positive illnesses but were markedly more common than in swab negative illnesses. The current UK definition had a sensitivity and specificity of 81% and 47% respectively for symptomatic COVID-19 compared to 93% and 26% for the broader definition. On average, cases met the broader case definition 0.3 days earlier than the current definition. 1.7-fold more illnesses met the broader definition than the current case definition.

**Interpretation:** COVID-19 is difficult to distinguish from other respiratory infections and common ailments on the basis of symptoms. Broadening the list of symptoms used to encourage engagement with TTI could moderately increase the number of infections identified and shorten delays to isolation, but with a large increase in the number of tests needed and the number of unwell individuals and contacts who are advised to self-isolate whilst awaiting results, and subsequently test negative for SARS-CoV-2.

## Introduction

The natural history of Severe acute respiratory syndrome virus-2 (SARS-CoV-2) infection can range from asymptomatic infection in around 25% of cases^1^ to severe or fatal Coronavirus disease-2019 (COVID-19) at a rate that is highly age dependent^2^. Understanding the natural history of symptomatic COVID-19 in the community is critical to the control of infection because it informs decisions about who should seek testing, whether those with symptoms should self-isolate, and whether those in contact with symptomatic people should self-isolate. Understanding the normal course of symptoms is also potentially helpful to patients and clinicians assessing whether care needs to be escalated due to unexpectedly severe or prolonged symptoms. In future, symptom profiles may also trigger early use of antivirals to prevent deterioration and potentially minimise transmission^3^. Finally, understanding symptom profiles is important to inform syndromic surveillance.

A wide range of clinical case definitions for COVID-19 are available utilising different combinations of symptoms to alert individuals to the need for testing, isolation and contact tracing. For example, WHO include the following symptoms in the clinical case definition of a suspected case: acute onset of fever AND cough; OR acute onset of ANY THREE OR MORE of the following signs or symptoms: fever, cough, general weakness/fatigue, headache, myalgia, sore throat, coryza, dyspnoea, anorexia/nausea/vomiting, diarrhoea, altered mental status^4^. European Centers for Disease Control define a possible case based on at least one of cough, fever, shortness of breath or sudden loss of sudden onset of anosmia, ageusia or dysgeusia^4^; and US Centers for Disease Control use the following clinical criteria: at least two of fever (measured or subjective), chills, rigors, myalgia, headache, sore throat, new olfactory and taste disorder(s) OR at least one of fever (measured or subjective), chills, rigors, myalgia, headache, sore throat, new olfactory and taste disorder(s)^5^.

In the UK, the Test Trace and Isolate community testing programme (TTI) asks individuals to seek testing if they have any of the following symptoms: a new continuous cough, a high temperature, loss of or altered sense of smell or taste^6^.

Although it is clear that addition of further symptoms could increase sensitivity, this is likely to be at the cost of reduced specificity and increasing numbers of people who require testing, isolation and contact tracing^7^.

For example, recent analysis of data from the REACT community survey suggests that adding in loss of appetite, chills, headache or muscle aches could increase the proportion of cases identified from 53% to 75% but at the cost of a 2.7-fold increase in required testing capacity^8^. Altering clinical case definitions will have different implications at varying levels of prevalence, since positive and negative predictive values of tests depend upon disease prevalence as well as test sensitivity and specificity. The timing within the course of an illness at which infected people meet the case definition is important since earlier isolation of cases and contacts could reduce transmission.

Prospective community studies where participants record symptoms in near-real time are needed to accurately measure COVID-19 symptom profiles with minimal recall bias. Here we describe prospectively recorded symptom profiles (frequency, severity and duration) of illnesses that tested positive for COVID-19 and illnesses that tested negative within a large community cohort study (Virus Watch). We compare the test characteristics (sensitivity, specificity, positive and negative predictive values, timing of meeting the case definition, number needed to test to identify one case) for the current UK definition and a wider definition proposed following analysis of the REACT study, which adds headache or chills or loss of appetite or muscle aches to the existing UK definition. We explore how symptom profiles and case definition performance vary by age and stage of the pandemic. We also publish the dataset from which these analyses were conducted allowing readers to explore the implications of different symptom combinations on case definition performance.

## Methods

### Study design and data collection

The Virus Watch study is a prospective, community cohort study which has been following entire households in England and Wales since 22 June 2020, over the COVID-19 pandemic. As of 04 May 2021, 24,296 households and 50,699 people across England and Wales have joined the study. The full study protocol is published elsewhere^9^.

In brief, participating households completed a baseline survey at enrollment into the study, followed by prospective, detailed daily symptom diaries recording the presence and severity of any symptoms of acute respiratory and gastrointestinal infections during any episodes of illness occurring during follow-up. At the end of each week of follow-up, households were emailed a survey link to report any symptoms from the preceding week as well as results of any SARS-CoV-2 swab tests conducted, including testing through TTI and routine asymptomatic screening, and including both polymerase chain reaction (PCR) and lateral flow device testing. Within the main cohort there was a nested sub-cohort of 10,766 participants who additionally provided study-specific swab specimens following predetermined symptom triggers (Appendix I) tested for SARS-CoV-2 by real-time reverse transcriptase PCR (RT-PCR) from the end of December 2020 onwards.

Symptom data gathered through the weekly survey were grouped into illness episodes and matched to swab results (see Appendix). The start date of an illness episode was defined as the first day any symptoms were reported, and the end date was the final day of reported symptoms. A 7-day washout period where no symptoms were reported was used to define the end of one illness episode and the start of a new illness episode. The data presented in this analysis includes illnesses which began between 22 June 2020 (start of the study) through to 02 May 2021. Within illness episodes, we investigated a wide range of individual symptoms (see Appendix) and the following symptom groupings: **UK Case definition** – one or more of the following: cough, measured fever (>=37.8C) or feeling feverish, or loss of, or altered, sense of smell or taste. **Broader case definition** -one or more of the following: cough, measured fever (>=37.8C) or feeling feverish, loss of, or altered, sense of smell or taste, headache, muscle aches, loss of appetite or chills.

### Analysis

We present simple descriptive analyses of the frequency, severity and duration of symptoms in SARS-CoV-2 test positive and test negative illnesses. We calculated sensitivity and specificity for individual symptoms and, for the two case definitions, we also calculated the positive predictive value (PPV), negative predictive value (NPV) number of people meeting the definition within the cohort and the numbers needed to test to identify one positive case (NNT) (Appendix II Figure S1).

We calculated the proportion of illnesses with SARS-CoV-2 test results and how this varied by UK case definition status and over time. We also calculated the ratio of the proportion of illnesses getting tested that met the UK case definition to those that did not meet the UK case definition. For both of these estimates we excluded illnesses beginning before 28 September 2020 as prior to this, our weekly surveys did not ask participants to report test results in weeks where there were no reported symptoms (Appendix I). This could have resulted in omission of some test results, particularly if there was a lag between swabbing and receipt of results. We also excluded illnesses beginning in the final week of data collection (week commencing 26 April 2021) as there may have been insufficient time to receive and report swab results.

### Data availability

We provide a dataset of illnesses and details of symptoms, swab test results, age group and time period of illness but other details removed to preserve anonymity. This can be used to assess sensitivity and specificity of different symptom combinations.

## Results

Overall, there were 29,083 illnesses (with at least one of the symptoms as defined in supplementary Table S1) reported in the cohort between 22 June 2020 and 02 May 2021. 8,213 (28.2%) illnesses were swabbed and of these 944 (11.5%) tested positive for SARS-CoV-2; 436 of these swabs were conducted as part of the study, including 22 positives. The percentage of swabbed illnesses testing positive were highest in young adults aged 16-24 years and lowest in children aged 0-15 years; highest in London and lowest in the South East and South West regions; and peaked in December 2020 (Table 1). Those with symptoms meeting the UK case definition for swabbing by TTI were more likely to be swabbed than those with other symptoms (Appendix II Table S2).

**Table 1.**
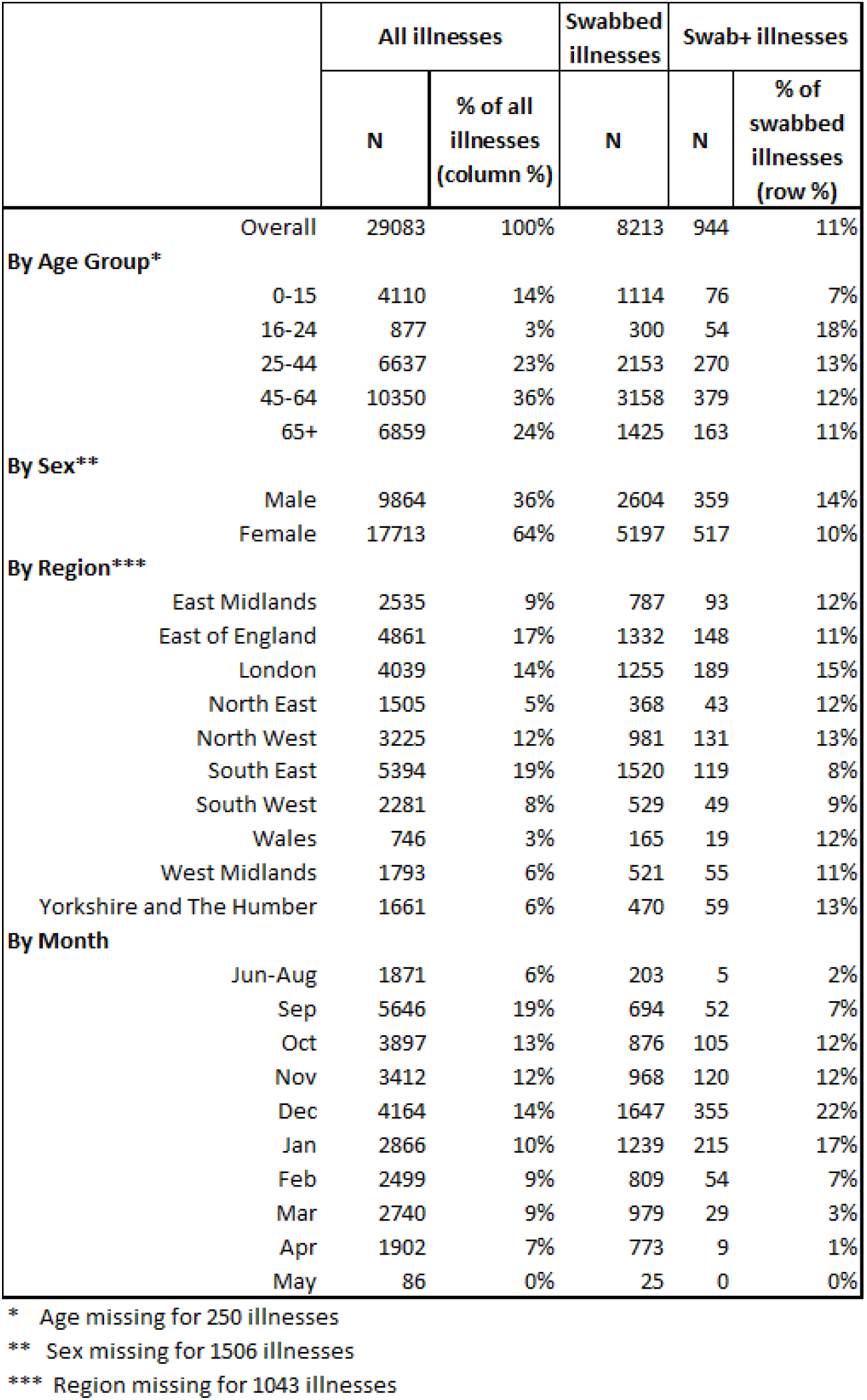
Characteristics of illnesses by demographics and swab outcome

|  | All illnesses |  | Swabbed illnesses | Swab+ illnesses |  |
| --- | --- | --- | --- | --- | --- |
|  | N | % of all illnesses (column %) | N | N | % of swabbed illnesses (row %) |
| Overall | 29083 | 100% | 8213 | 944 | 11% |
| **By Age Group\*** |  |  |  |  |  |
| 0-15 | 4110 | 14% | 1114 | 76 | 7% |
| 16-24 | 877 | 3% | 300 | 54 | 18% |
| 25-44 | 6637 | 23% | 2153 | 270 | 13% |
| 45-64 | 10350 | 36% | 3158 | 379 | 12% |
| 65+ | 6859 | 24% | 1425 | 163 | 11% |
| **By Sex\*\*** |  |  |  |  |  |
| Male | 9864 | 36% | 2604 | 359 | 14% |
| Female | 17713 | 64% | 5197 | 517 | 10% |
| **By Region\*\*\*** |  |  |  |  |  |
| East Midlands | 2535 | 9% | 787 | 93 | 12% |
| East of England | 4861 | 17% | 1332 | 148 | 11% |
| London | 4039 | 14% | 1255 | 189 | 15% |
| North East | 1505 | 5% | 368 | 43 | 12% |
| North West | 3225 | 12% | 981 | 131 | 13% |
| South East | 5394 | 19% | 1520 | 119 | 8% |
| South West | 2281 | 8% | 529 | 49 | 9% |
| Wales | 746 | 3% | 165 | 19 | 12% |
| West Midlands | 1793 | 6% | 521 | 55 | 11% |
| Yorkshire and The Humber | 1661 | 6% | 470 | 59 | 13% |
| **By Month** |  |  |  |  |  |
| Jun-Aug | 1871 | 6% | 203 | 5 | 2% |
| Sep | 5646 | 19% | 694 | 52 | 7% |
| Oct | 3897 | 13% | 876 | 105 | 12% |
| Nov | 3412 | 12% | 968 | 120 | 12% |
| Dec | 4164 | 14% | 1647 | 355 | 22% |
| Jan | 2866 | 10% | 1239 | 215 | 17% |
| Feb | 2499 | 9% | 809 | 54 | 7% |
| Mar | 2740 | 9% | 979 | 29 | 3% |
| Apr | 1902 | 7% | 773 | 9 | 1% |
| May | 86 | 0% | 25 | 0 | 0% |

\* Age missing for 250 illnesses
\*\* Sex missing for 1506 illnesses
\*\*\* Region missing for 1043 illnesses

When excluding illnesses without full opportunity to report swab results, the proportion of illnesses reporting swab results steadily increases from September 2020 to January 2021, temporarily drops in February and increases again through April 2021 (Appendix II Figure S2). Illnesses meeting the case definition were more likely to be swabbed than illnesses not meeting the case definition, although the differences in swabbing behaviour of these two groups changed over time (Appendix II Figures S2 & S3). The ratio of the proportion of illnesses reporting a test result among those meeting the case definition to those not meeting the case definition was highest in September 2020, when those meeting the case definition were 4.4 times more likely to report a test result. This ratio steadily declined over time and by April 2021 illnesses meeting the case definition were only 1.3 more likely to report a test result compared to illnesses not meeting the case definition.

Figure 1a shows the proportion of swabbed illnesses that reported each symptom according to whether they tested positive or negative for SARS-CoV-2. All of the wide range of reported symptoms were more common in illnesses that tested positive than illnesses that tested negative. Amongst SARS-CoV-2-positive illnesses, the ten most commonly reported symptoms, in decreasing order of frequency, were: fatigue, headache, cough, muscle ache, needing to spend extra time in bed, loss of, or altered, sense of smell or taste, loss of appetite, sore throat, difficulties in undertaking daily activities, and bone or joint aches. The percentage of illnesses with each symptom by day of illness is shown in Figures 1b and c, which illustrates both the higher frequency and longer duration of key symptoms in test positive and test negative cases.

**Figure 1.**
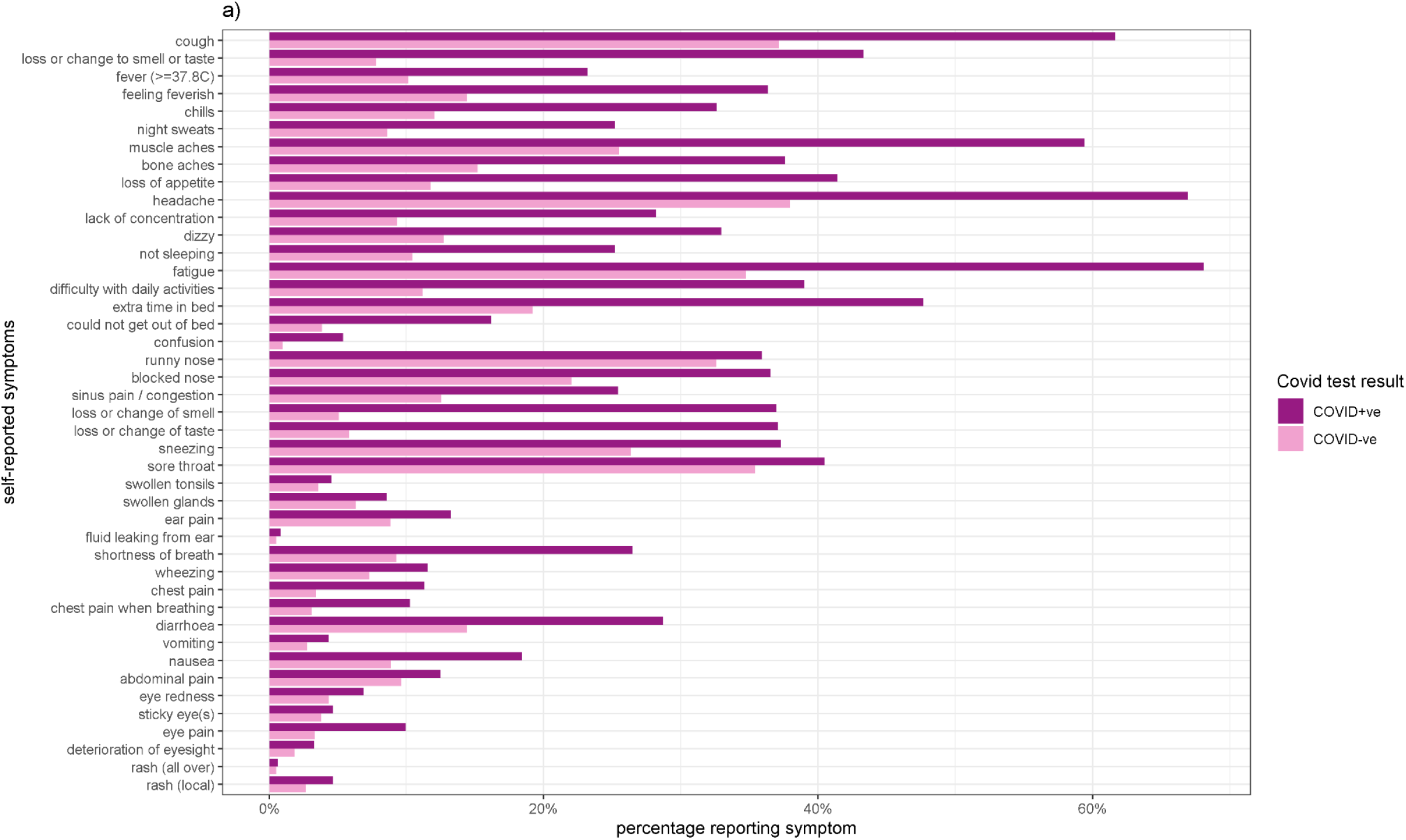
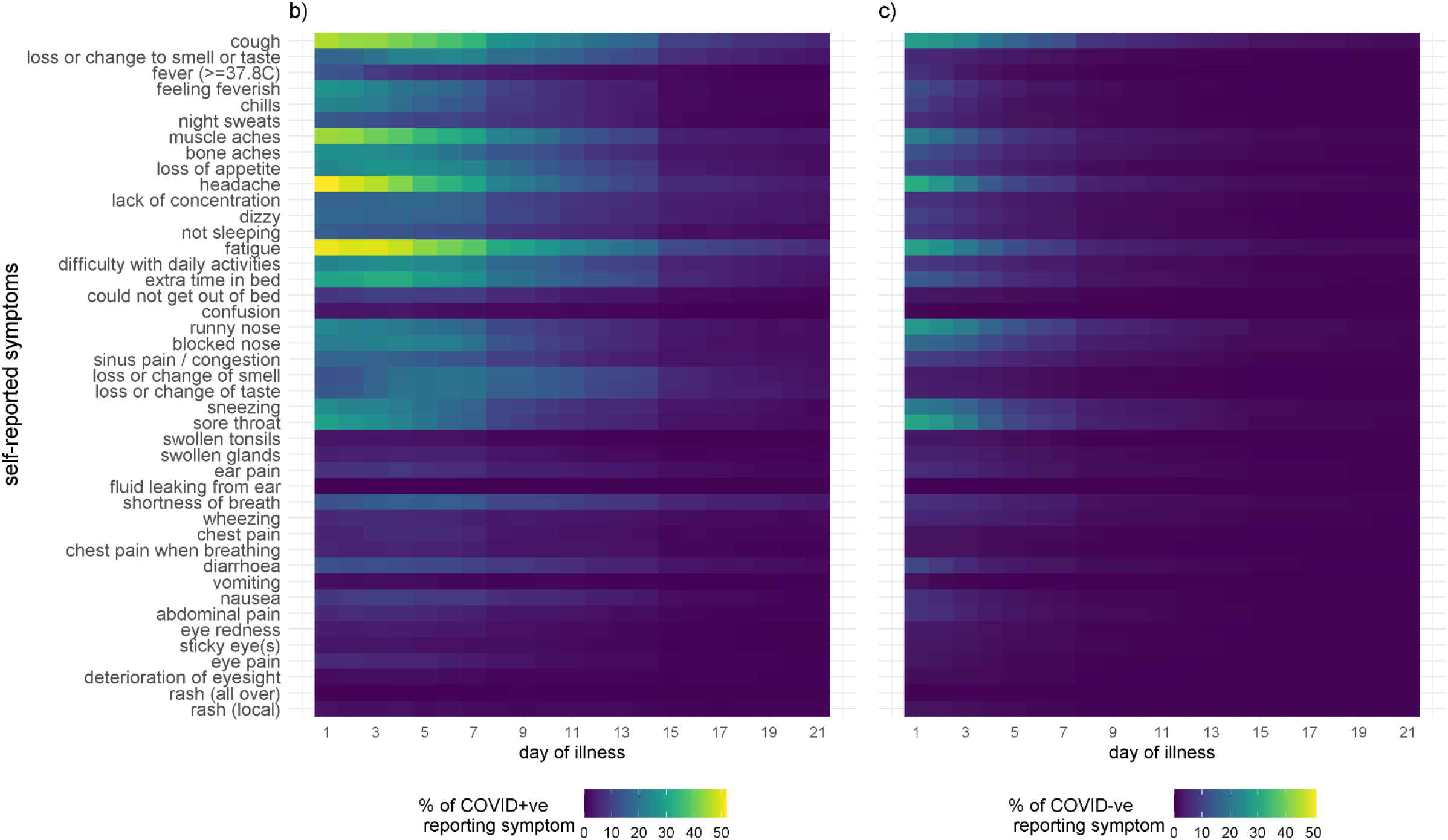
COVID-19 symptoms. **a**, Self-reported symptoms by swab-confirmed test positive and test negative illnesses. **b-c**, Proportion of COVID-19 illnesses **(b)** and non-COVID-19 illnesses **(c)** experiencing symptoms on a given day of illness within the first three week of illness. Day 1 represents the onset of symptoms.

Table S2 in the supplementary appendix shows the sensitivity and specificity of each symptom and the mean and median day of illness on which they are first reported among all swabbed illnesses with a reported result. Most symptoms arise early in the course of illness (mean onset typically between 2 to 2.5 days) although loss or change to sense of smell or taste and the two chest pain symptoms appear slightly later (day 3 to 3.5) during the course of illness. It can be seen that although cough and some constitutional symptoms such as headache, fatigue and muscle aches are fairly common among COVID-19 illnesses (i.e., they have a reasonably high sensitivity) they are also a common feature of non-COVID-19 illnesses and thus have a low corresponding specificity. In contrast the loss or change to sense of smell or taste is slightly less common among COVID-19 illnesses but comparatively much less common among non-COVID-19 illnesses, leading to a lower sensitivity but higher specificity.

Figure 2 and Table S3 (in the supplementary appendix) show the maximum reported severity for a range of key symptoms in test positive and test negative illnesses. When symptoms do occur, they are more likely to be severe in COVID-19 illnesses than in non-COVID-19 illnesses. Figure 3 shows the distribution of the duration of illnesses in test positive and test negative illnesses. It can be seen that the duration of illness is longer in COVID-19 illnesses than in other illnesses. Table S4 (in the supplementary appendix) shows how illness duration varies by age and gender in COVID-19 positive and negative illnesses and Figure S4 shows the distribution of duration of illnesses among COVID-19 illnesses by age group. Illnesses tend to be of longer duration in older individuals.

**Figure 2.**
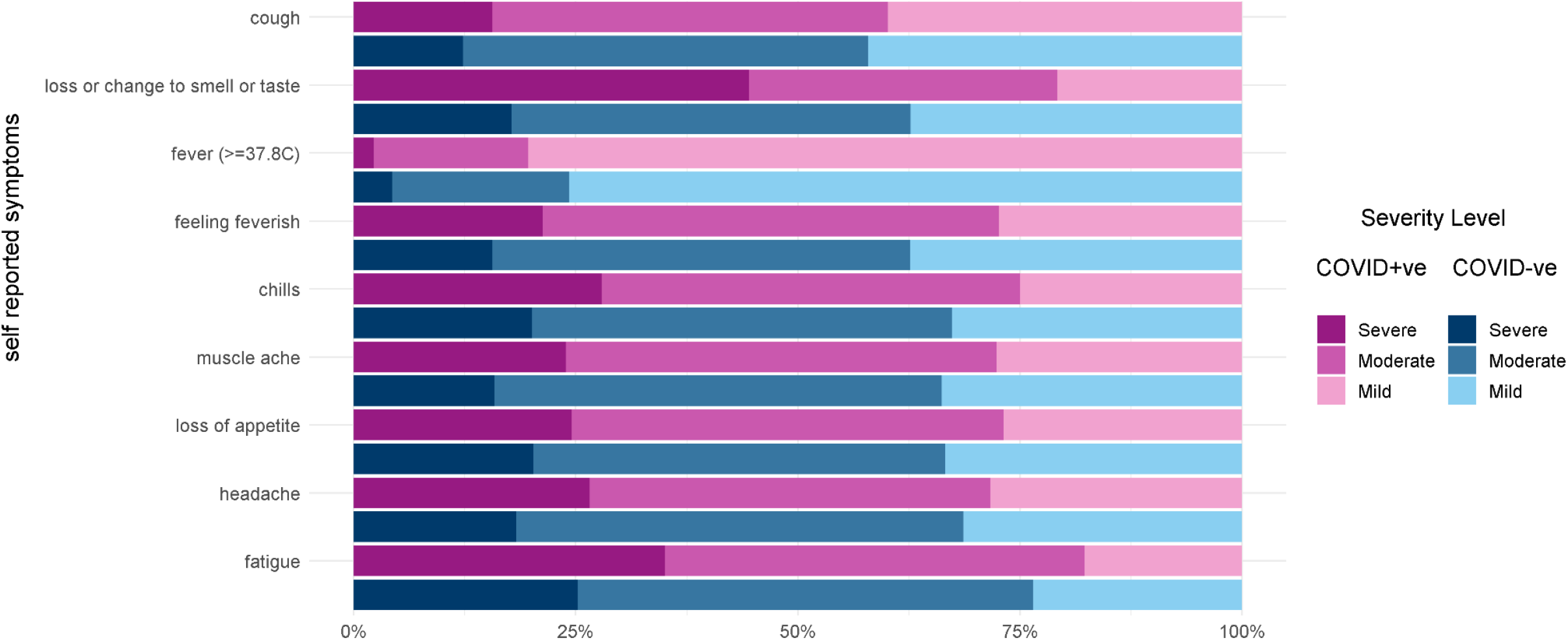
Severity of symptoms among swab-confirmed test positive and test negative illnesses reporting the symptom.

**Figure 3.**
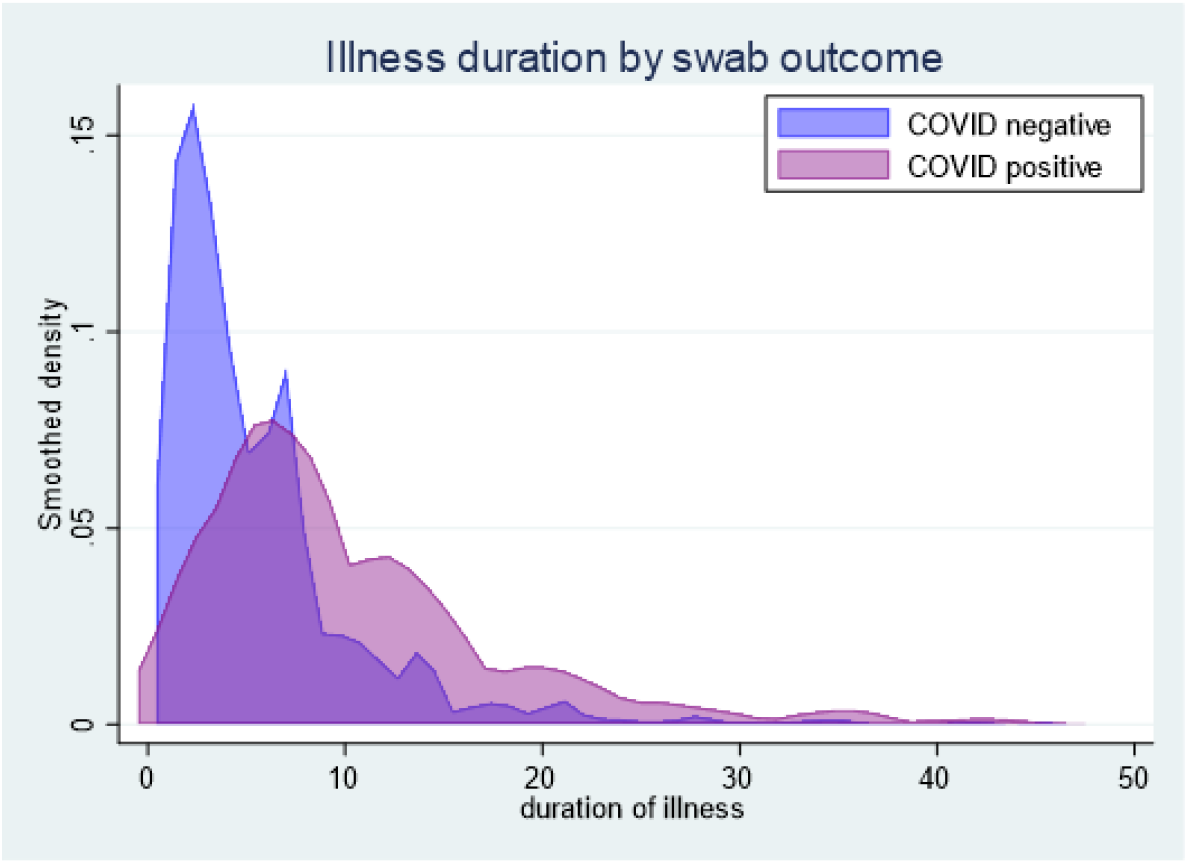
Distribution of illness duration* by COVID-19 status ^*^limited to illnesses with duration <50 days.

Table 2 shows the mean and median day of meeting the current and broader case definition, sensitivity, specificity, PPV, NPV, and NNT to identify a case. The numbers meeting the case definition and how many folds higher this is for the broader case definition are shown (multiplication factor). These results are stratified by age and calendar time.

**Table 2.**
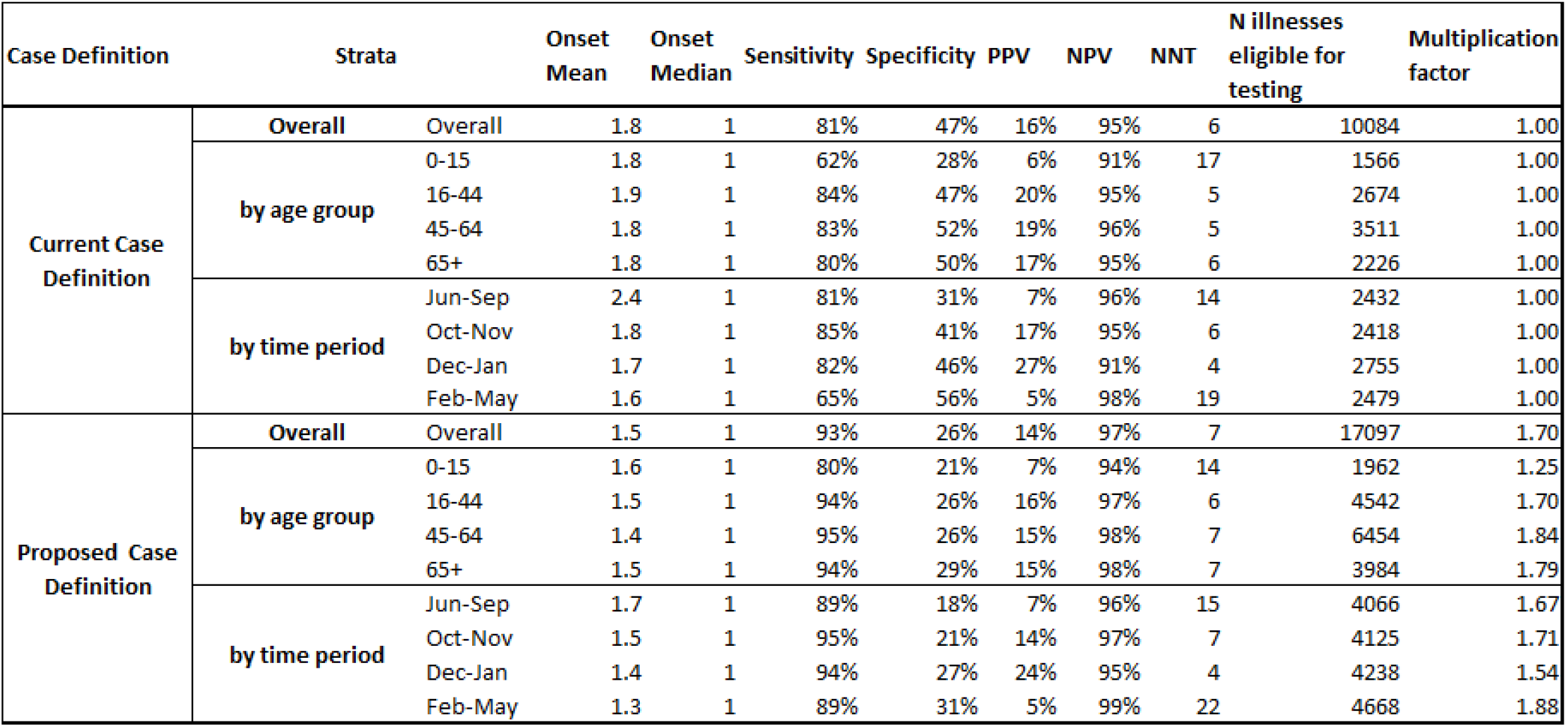
Speed of identifying cases, proportion of all community illnesses requiring testing and test characteristics for the current and proposed UK case definitions.

Sensitivity of the current UK case definition (i.e., the proportion of all illnesses testing positive who met the definition) was 81% compared to 93% for the broader case definition. Specificity (i.e. the proportion of all those illnesses testing negative who did not meet the case definition) was 47% for the current case definition and 26% for the broader case definition. Sensitivity and specificity of both case definitions was lower in children aged 0-15 years than in older age groups, particularly for the current case definition. Sensitivity of the current case definitions remained stable (81-85%) between June 2020 through January 2021 but decreased in February to May 2021 (65%). In contrast the sensitivity of the proposed case definition remained stable over the entire period (89-95%). Specificity of both case definitions increased over time. The PPV (the proportion of those meeting the clinical case definition who test positive) was 16% for the current UK case definition and 14% for the broader definition. PPV estimates were substantially lower (between 5-7%) in the youngest age group and during August through September 2020 and in February through May 2021, corresponding with the age group and time periods with the lowest disease rates. NPV (the proportion of those with illnesses not meeting the clinical case definition who tested negative) was 95% for the current UK case definition and 97% for the broader case definition. The number of illnesses meeting the broader case definition was 1.7-fold higher than those meeting the current definition.

## Discussion

We characterised the symptom profiles and estimated the accuracy of clinical case definitions for COVID-19 among community cases arising in a large, prospective, population-based cohort study based in the UK. All symptoms asked about were more frequently reported, and when present were generally more severe and longer-lasting, in COVID-19 illnesses compared to non-COVID-19 illnesses. Individually, cough and some constitutional symptoms including headache, muscle ache and fatigue presented early in the course of illness and had moderate sensitivity and specificity as they were common in both test positive and test negative illnesses. In contrast, fever and loss of or altered sense of smell or taste presented slightly later in illness and had a lower sensitivity but higher specificity as they were not as common in COVID-19 illnesses but were even less common in non-COVID-19 illnesses. The combination of symptoms in the current UK case definition had a higher sensitivity (81%) than any individual symptom, with a specificity of 47%. Adding additional symptoms to the case definition could lead to earlier case identification and higher sensitivity, but at the cost of specificity and consequently, a substantial increase in the number of non-COVID-19 illnesses eligible for testing. For example, when we compare the broader case definition to the current UK case definition, cases on average met the case definition 0.3 days earlier and there was a moderate increase in sensitivity to 93% but at a much lower specificity of 26%, with 1.7 times more illnesses eligible for testing.

Strengths of this work include the prospective daily recording of a wide range of symptoms across a large community cohort and linkage of these to self-reported swab results ascertained on a weekly basis. This should maximise the accuracy of symptom data amongst swabbed participants. While not fully representative of the population of England and Wales, our sample includes participants in every local authority area; however, there is a moderate overrepresentation of those aged over 65 and an underrepresentation of those in more deprived areas^10^. Whilst we collected information on a very wide range of symptoms, testing primarily relied on that conducted through the national TTI programme meaning that those meeting the current case definition are more likely to be tested. This is likely to lead to an overestimation of the sensitivity of the current UK case definition. This bias towards testing illnesses meeting the case definition however was reduced over time. This change in testing behaviour and the drop in sensitivity seen between February-May 2021 is likely to be attributed, at least in part, to the roll-out and widespread availability of at-home lateral flow testing to the wider population from March 2021. The availability and ease of at-home lateral flow devices may have lowered individuals’ threshold for testing, leading to more mild illnesses being tested. A further strength is that we have published the dataset online to enable replication of analyses and for others to explore the test characteristics of various combinations of symptoms for case definitions.

COVID-19 is difficult to distinguish from other respiratory infections or common ailments on the basis of symptoms alone. Also, a high proportion of infections are asymptomatic or have a pre-symptomatic phase when transmission can occur. As such, systems to identify symptomatic cases, test them, and isolate cases and their contacts can only ever reduce, rather than prevent all transmission. These programmes, when part of a broader programme of non-pharmaceutical interventions, can however contribute to control of infection and may be particularly effective when introduced during periods of lower incidence. For example, countries that combined early and strict border controls, intensive testing, and rapid triggering of lockdowns have had substantially lower COVID-19 transmission and mortality than other countries such as the UK^11^.

The success of testing and isolation programmes is dependent on public understanding and engagement. Data from behavioural surveys in England show only 51% of participants knew the symptoms that testing is recommended for, only 18% sought testing if they had the symptoms and only 42.5% fully adhered to self-isolation. Engagement was lower in younger people and amongst those in financial hardship^12^. Engagement with population level asymptomatic testing using lateral flow testing has also been shown to be low, particularly in socioeconomically disadvantaged areas^13^.

Policy makers considering which symptoms might prompt testing, tracing and isolation need to balance the availability of testing capacity at different stages of the pandemic, the speed with which samples can be taken and results returned, the harms incurred by asking large numbers of people who do not have COVID-19 and their contacts to self-isolate whilst awaiting test results, the consistency and simplicity of public health messaging and the likely public engagement with the system. Alteration of symptom profiles triggering testing and isolation may have less impact than other approaches to increase uptake and engagement, and to ensure timely and effective tracing of household and non-household contacts.

The fact that COVID-19 may present as any of a very wide range of symptoms, or with no symptoms at all, is one of the key challenges in implementing successful TTI systems. Low levels of engagement also limit effectiveness. This emphasises the importance of not placing undue reliance on such systems as a mechanism to allow relaxation of other social distancing measures and the critical importance of protecting populations globally through immunisation.

## Supporting information

Fragaszy_et_al_Symptom_Profiles_Supplemental_Appendix

## Data Availability

We aim to share aggregate data from this project on our website and via a "Findings so far" section on our website - https://ucl-virus-watch.net/. We will also be sharing individual record level data on a research data sharing service such as the Office of National Statistics Secure Research Service. In sharing the data we will work within the principles set out in the UKRI Guidance on best practice in the management of research data. Access to use of the data whilst research is being conducted will be managed by the Chief Investigators (ACH and RWA) in accordance with the principles set out in the UKRI guidance on best practice in the management of research data. We will put analysis code on publicly available repositories to enable their reuse.

## References

1. Alene M, Yismaw L, Assemie MA, et al. Magnitude of asymptomatic COVID-19 cases throughout the course of infection: A systematic review and meta-analysis. Kwok KO, ed. PLoS ONE. 2021;16(3):e0249090. doi:10.1371/journal.pone.0249090

2. Levin AT, Hanage WP, Owusu-Boaitey N, Cochran KB, Walsh SP, Meyerowitz-Katz G. Assessing the age specificity of infection fatality rates for COVID-19: systematic review, meta-analysis, and public policy implications. Eur J Epidemiol. 2020;35(12):1123–1138. doi:10.1007/s10654-020-00698-1

3. Government launches COVID-19 Antivirals Taskforce to roll out innovative home treatments this autumn. GOV.UK. Accessed May 13, 2021. https://www.gov.uk/government/news/government-launches-covid-19-antivirals-taskforce-to-roll-out-innovative-home-treatments-this-autumn

4. WHO COVID-19 Case definition. Accessed May 13, 2021. https://www.who.int/publications-detail-redirect/WHO-2019-nCoV-Surveillance_Case_Definition-2020.2

5. Coronavirus Disease 2019 (COVID-19) | 2020 Interim Case Definition, Approved April 5, 2020. Accessed May 13, 2021. /nndss/conditions/coronavirus-disease-2019-covid-19/case-definition/2020/

6. Get a free PCR test to check if you have coronavirus (COVID-19). GOV.UK. Accessed May 13, 2021. https://www.gov.uk/get-coronavirus-test

7. Antonelli M, Capdevila J, Chaudhari A, et al. Optimal symptom combinations to aid COVID-19 case identification: analysis from a community-based, prospective, observational cohort. medRxiv. Published online February 8, 2021:2020.11.23.20237313. doi:10.1101/2020.11.23.20237313

8. Elliott J, Whitaker M, Bodinier B, et al. Symptom reporting in over 1 million people: community detection of COVID-19. medRxiv. Published online February 12, 2021:2021.02.10.21251480. doi:10.1101/2021.02.10.21251480

9. Hayward A, Fragaszy E, Kovar J, et al. Risk factors, symptom reporting, healthcare-seeking behaviour and adherence to public health guidance: protocol for Virus Watch, a prospective community cohort study. medRxiv. Published online December 16, 2020:2020.12.15.20248254. doi:10.1101/2020.12.15.20248254

10. UCL Virus Watch - help stop the spread of COVID-19. Accessed May 13, 2021. https://ucl-virus-watch.net

11. Oliu-Barton M, Pradelski BSR, Aghion P, et al. SARS-CoV-2 elimination, not mitigation, creates best outcomes for health, the economy, and civil liberties. The Lancet. 2021;0(0). doi:10.1016/S0140-6736(21)00978-8

12. Smith LE, Potts HWW, Amlôt R, Fear NT, Michie S, Rubin GJ. Adherence to the test, trace, and isolate system in the UK: results from 37 nationally representative surveys. BMJ. 2021;372:608. doi:10.1136/bmj.n608

13. Liverpool COVID-19 community testing pilot: interim evaluation report summary. GOV.UK. Accessed May 13, 2021. https://www.gov.uk/government/publications/liverpool-covid-19-community-testing-pilot-interim-evaluation-report-summary/liverpool-covid-19-community-testing-pilot-interim-evaluation-report-summary

