## Supplementary material for "Symptom profiles and accuracy of clinical definitions for COVID-19 in the community. Results of the Virus Watch community cohort": Fragaszy_et_al_Symptom_Profiles_Supplemental_Appendix

#### I. Additional Methods

##### Reporting of swab tests taken outside the study

Throughout the study, participants reporting symptoms in the previous week were also asked to report the outcome of any NHS COVID-19 swab test for that illness. From the week commencing 28 September 2020, participants could also report the results of any swab testing in the previous week, regardless of whether they were also reporting symptoms for that week. This was to ensure that all swab test results could be reported regardless of when the test results arrived or whether the swab was taken asymptomatically.

##### Tests taken within the study

The laboratory sub-cohort is split into three groups with different swabbing protocols. In all three groups, participants are asked to provide self-administered swabs on day two of illness if they experience one or more of the trigger symptoms. Group 1 (7,368 participants) swab for respiratory symptoms, fever or feverishness or loss or change to sense of smell or taste. Group 2 (1,042 participants) swab for respiratory, gastrointestinal or general symptoms of infection including fever or feverishness or loss or change to sense of smell or taste. Group 3 (2,216 participants) swab for cough, fever or feverishness or loss or change to sense of smell or taste. In group 3, if any household member experiences a trigger symptom then all household contacts are also asked to swab on the same day as the index case regardless of whether they are symptomatic and if any of those swabs are PCR+ then all household members are asked to swab again one and two weeks later irrespective of symptoms.

#### Matching of illness episodes and test results

To differentiate COVID-19 from non-COVID-19 illnesses, we matched illness episodes and self-reported swab test results that were within up to two weeks of each other. Matching was undertaken in a stepwise manner, for both symptomatic swabbing and asymptomatic swabbing, on a week-by-week basis, based on the likely sequence of symptoms and swab positivity. First, swab test results were matched to illnesses occurring in the same week as receipt of the swab result. Following that, any test results that did not link to an illness in the same week, were sequentially matched to illness episodes that had commenced in the following week (+1 week), then finished in the week prior (-1 week), and then finished two weeks prior (-2 weeks). Illnesses commencing 2 weeks after a swab result were not matched as these were considered unlikely to be related. For each round of sequential matching, only test results not yet matched to an illness were used.

Some illness episodes continued for several weeks, and the associated swab test may have been completed any time during the episode, or shortly before or after it. If more than one swab test result matched to an illness episode, any positive swab test result superseded any negative swab test result. Any illness episode matching to a positive swab result was taken to be a confirmed COVID-19 illness. Any illness episode matching to negative swab test results only, was taken to be a non-COVID-19 illness. Illness episodes not matching any swab test results, and swab test results not matching to any illness episodes, were not included in these analyses. Where an individual had multiple test positive illnesses, we only analysed their first test positive illness as it was felt that this was most likely to be the result of long-term virus shedding rather than re-infection.

#### Calculation of test characteristics for clinical case definitions

We calculated the diagnostic test characteristics of clinical case definitions and individual symptoms using the formulae from Figure S1.

### II. Additional Tables and Figures

Figure S1. Two by two contingency table and equations used to calculate test characteristics of the clinical case definitions compared to swab test results (gold standard).

| | | Gold standard test (Swab test result) Positive | Gold standard test (Swab test result) Negative | Totals |
|---|---|---|---|---|
| Clinical Case Definition | Positive | a | b | a+b |
| | Negative | c | d | c+d |
| Totals | | a+c | b+d | a+b+c+d |

| | |
|---|---|
| True Positives = a | False Positives = b |
| True Negatives = d | False Negatives = c |

$$True\ prevalence = \frac{a + c}{a + b + c + d}$$

$$Sensitivity = \frac{a}{a + c}$$

$$Specificity = \frac{d}{b + d}$$

$$Predictive\ value\ of\ a\ positive\ test\ (PPV) = \frac{a}{a + b}$$

$$Predictive\ value\ of\ a\ negative\ test\ (NPV) = \frac{d}{c + d}$$

$$Number\ needed\ to\ test\ (NNT) = \frac{1}{PPV}$$

Figure S2. Percent of illnesses with a swab test result by month, overall and by UK Case Definition status

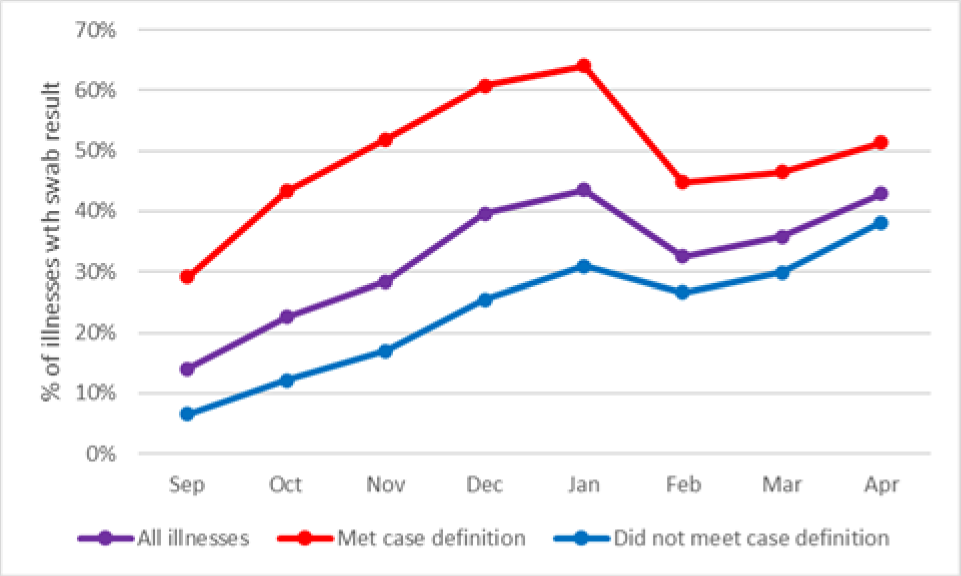


Figure S3. Ratio of swabbing among illnesses meeting the case definition to those not meeting the case definition over time.

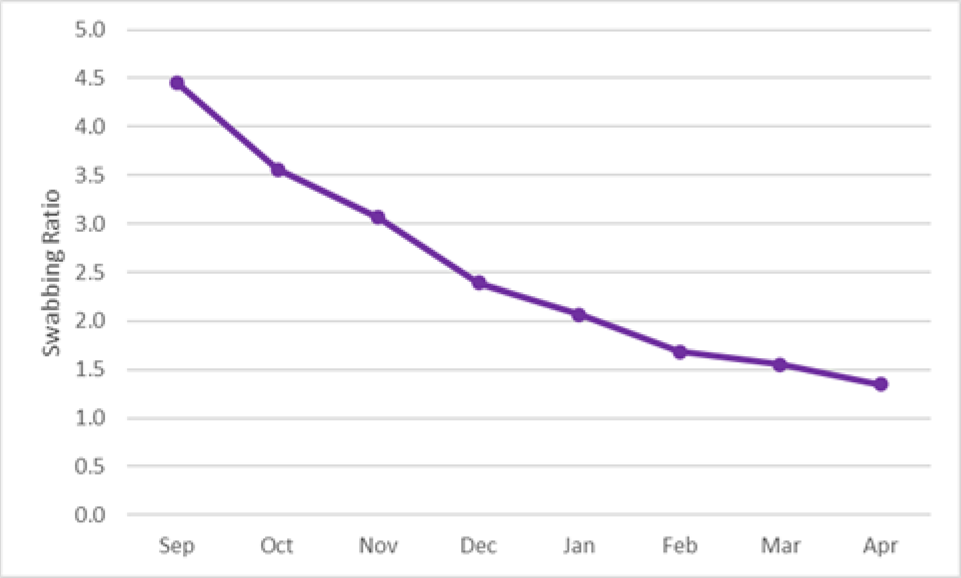

Table S1. Details of symptoms and symptom groupings

| | Symptom name | Symptom wording and combinations |
|---|---|---|
| **Grouped Symptoms (used in case defintions)** | cough | This was a combination of the following individual symptoms: 'dry_cough', 'white_phlegm' and 'green_phlegm' (i.e. "Dry Cough" AND/OR "Coughing up White Phlegm" AND/OR "Coughing up Green Phlegm") |
| | fever | This was a combination of the following individual symptoms: 'fever_cat' and 'feeling_feverish' (i.e. "Fever" with reported temperature >=37.8C AND/OR "Feeling feverish") |
| | smell_taste | This was a combination of the following individual symptoms: 'loss_of_smell' and 'loss_of_taste' (i.e. "Loss or change to sense of smell" AND/OR "Loss or change to sense of taste") |
| **Individual Symptoms** | fever_cat | "Fever" with reported temperature >=37.8C |
| | feeling_feverish | "Feeling Feverish" |
| | chills | "Chills and shakes" |
| | night_sweats | "Night Sweats" |
| | muscle_ache | "Muscle Ache" |
| | bone_ache | "Bone or Joint ache" |
| | loss_of_appetite | "Loss of Appetite" |
| | headache | "Headache" |
| | concentration | "Lack of Concentration" |
| | dizzy | "Lightheaded or Dizzy" |
| | not_sleeping | "Not Sleeping" |
| | fatigue | "Fatigue / Feeling Unusually Tired" |
| | daily_activities | "Difficulties with daily activities around the house" |
| | extra_bed | "Needed extra time in bed" |
| | out_of_bed | "Could not get out of bed" |
| | confusion | "Confusion, disorientation, or hallucinations (altered mental state)" |
| | runny_nose | "Runny Nose" |
| | blocked_nose | "Blocked Nose" |
| | sinus_pain | "Sinus Pain / congestion" |
| | dry_cough | "Dry cough" |
| | white_phlegm | "Coughing up White Phlegm" |
| | green_phlegm | "Coughing up Green Phlegm" |
| | loss_of_smell | "Loss or change to sense of smell" |
| | loss_of_taste | "Loss or change to sense of taste" |
| | sneezing | "Sneezing" |
| | sore_throat | "Sore Throat" |
| | swollen_tonsils | "Swollen Tonsils" |
| | swollen_glands | "Swollen Glands (anlarged lymph nodes)" |
| | ear_pain | "Ear Pain or change in hearing" |
| | ear_fluid | "Fluid leaking from ear" |
| | sob | "Shortness of Breath / difficulty breathing" |
| | wheezing | "Wheezing" |
| | chest_pain_nc | "Chest pain (not changed by breathing or moving)" |
| | chest_pain_br | "Chest pain when breathing in" |
| | diarrhoea | "Diarrhoea (even mild)" |
| | vomiting | "Vomiting (being sick)" |
| | nausea | "Nausea (feeling sick) |
| | abdo_pain | "Abdominal pain (not including menstrual pain)" |
| | redeye | "Eye Redness" |
| | sticky_eye | "Sticky Eye(s)" |
| | eye_pain | "Eye Pain" |
| | eye_deter | "Deterioration of eyesight" |
| | rash_allover | "Rash - All Over" |
| | rash_local | "Rash - Local" |

Table S2. Speed of symptom onset, proportion of all community cases tested, and the test characteristics of each symptom.

| Symptom | Onset Mean | Onset Median | Sensitivity | Specificity | PPV | NPV | Proportion tested* |
|---|---|---|---|---|---|---|---|
| cough_bin | 2.0 | 1 | 62% | 63% | 18% | 93% | 53% |
| smell_taste_bin | 3.0 | 1 | 43% | 92% | 42% | 93% | 77% |
| fever_bin | 1.9 | 1 | 45% | 79% | 22% | 92% | 62% |
| fevercat_bin | 2.1 | 1 | 23% | 90% | 23% | 90% | 78% |
| feeling_feverish_bin | 1.9 | 1 | 36% | 86% | 25% | 91% | 58% |
| chills_bin | 2.0 | 1 | 33% | 88% | 26% | 91% | 53% |
| night_sweats_bin | 2.4 | 1 | 25% | 91% | 28% | 90% | 51% |
| muscle_ache_bin | 1.8 | 1 | 59% | 74% | 23% | 93% | 44% |
| bone_ache_bin | 2.0 | 1 | 38% | 85% | 24% | 91% | 41% |
| loss_of_appetite_bin | 2.3 | 1 | 41% | 88% | 31% | 92% | 59% |
| headache_bin | 1.8 | 1 | 67% | 62% | 19% | 94% | 41% |
| concentration_bin | 2.2 | 1 | 28% | 91% | 28% | 91% | 48% |
| dizzy_bin | 2.5 | 1 | 33% | 87% | 25% | 91% | 48% |
| not_sleeping_bin | 2.4 | 1 | 25% | 90% | 24% | 90% | 42% |
| fatigue_bin | 1.7 | 1 | 68% | 65% | 20% | 94% | 44% |
| daily_activities_bin | 2.3 | 1 | 39% | 89% | 31% | 92% | 51% |
| extra_bed_bin | 2.2 | 1 | 48% | 81% | 24% | 92% | 48% |
| out_of_bed_bin | 2.5 | 1 | 16% | 96% | 35% | 90% | 55% |
| confusion_bin | 3.1 | 1 | 5% | 99% | 41% | 89% | 55% |
| runny_nose_bin | 1.7 | 1 | 36% | 67% | 13% | 89% | 33% |
| blocked_nose_bin | 2.0 | 1 | 37% | 78% | 18% | 90% | 37% |
| sinus_pain_bin | 2.3 | 1 | 25% | 87% | 21% | 90% | 43% |
| dry_cough_bin | 2.2 | 1 | 53% | 73% | 20% | 92% | 59% |
| white_phlegm_bin | 3.0 | 1 | 18% | 90% | 19% | 89% | 44% |
| green_phlegm_bin | 3.3 | 1 | 8% | 94% | 15% | 89% | 51% |
| loss_of_smell_bin | 3.0 | 1 | 37% | 95% | 49% | 92% | 81% |
| loss_of_taste_bin | 3.1 | 1 | 37% | 94% | 45% | 92% | 79% |
| sneezing_bin | 1.9 | 1 | 37% | 74% | 16% | 90% | 34% |
| sore_throat_bin | 1.7 | 1 | 40% | 65% | 13% | 89% | 41% |
| swollen_tonsils_bin | 2.3 | 1 | 5% | 96% | 14% | 89% | 54% |
| swollen_glands_bin | 2.7 | 1 | 9% | 94% | 15% | 89% | 51% |
| ear_pain_bin | 2.6 | 1 | 13% | 91% | 16% | 89% | 41% |
| ear_fluid_bin | 4.1 | 1 | 1% | 99% | 18% | 89% | 36% |
| sob_bin | 2.7 | 1 | 26% | 91% | 27% | 90% | 56% |
| wheezing_bin | 2.8 | 1 | 12% | 93% | 17% | 89% | 52% |
| chest_pain_nc_bin | 3.3 | 1 | 11% | 97% | 30% | 89% | 60% |
| chest_pain_br_bin | 3.5 | 1 | 10% | 97% | 30% | 89% | 68% |
| diarrhoea_bin | 2.1 | 1 | 29% | 86% | 21% | 90% | 32% |
| vomiting_bin | 2.6 | 1 | 4% | 97% | 17% | 89% | 34% |
| nausea_bin | 2.2 | 1 | 18% | 91% | 21% | 90% | 37% |
| abdo_pain_bin | 2.3 | 1 | 13% | 90% | 14% | 89% | 33% |
| redeye_bin | 2.4 | 1 | 7% | 96% | 17% | 89% | 32% |
| sticky_eye_bin | 2.3 | 1 | 5% | 96% | 14% | 89% | 29% |
| eye_pain_bin | 2.7 | 1 | 10% | 97% | 28% | 89% | 38% |
| eye_deter_bin | 2.7 | 1 | 3% | 98% | 19% | 89% | 33% |
| rash_allover_bin | 2.5 | 1 | 1% | 99% | 14% | 89% | 36% |
| rash_local_bin | 2.6 | 1 | 5% | 97% | 18% | 89% | 33% |

* Includes tested illnesses where test result was not reported to the study

Table S3. Severity of symptoms among swab-confirmed COVID-19 positive and negative illnesses.

| Symptoms | COVID+ illnesses (N=944) | | | | | COVID- illnesses (N=7269) | | | | |
|---|---|---|---|---|---|---|---|---|---|---|
| | n | % | Max severity during illness (when symptom is present) | | | n | % | Max severity during illness (when symptom is present) | | |
| | | | % mild | % mod | % severe | | | % mild | % mod | % severe |
| cough | 582 | 62% | 40% | 45% | 16% | 2699 | 37% | 42% | 46% | 12% |
| smell_taste | 409 | 43% | 21% | 35% | 44% | 568 | 8% | 37% | 45% | 18% |
| fevercat | 219 | 23% | 80% | 17% | 2% | 738 | 10% | 76% | 20% | 4% |
| feeling_feverish | 343 | 36% | 27% | 51% | 21% | 1049 | 14% | 37% | 47% | 16% |
| chills | 308 | 33% | 25% | 47% | 28% | 876 | 12% | 33% | 47% | 20% |
| night_sweats | 238 | 25% | 26% | 40% | 34% | 626 | 9% | 29% | 49% | 22% |
| muscle_ache | 561 | 59% | 28% | 48% | 24% | 1855 | 26% | 34% | 50% | 16% |
| bone_ache | 355 | 38% | 27% | 46% | 27% | 1105 | 15% | 30% | 49% | 21% |
| loss_of_appetite | 391 | 41% | 27% | 49% | 25% | 855 | 12% | 33% | 46% | 20% |
| headache | 632 | 67% | 28% | 45% | 27% | 2759 | 38% | 31% | 50% | 18% |
| concentration | 266 | 28% | 38% | 46% | 15% | 678 | 9% | 37% | 46% | 17% |
| dizzy | 311 | 33% | 43% | 39% | 18% | 925 | 13% | 46% | 39% | 15% |
| not_sleeping | 238 | 25% | 28% | 50% | 22% | 758 | 10% | 32% | 47% | 21% |
| fatigue | 643 | 68% | 18% | 47% | 35% | 2527 | 35% | 24% | 51% | 25% |
| daily_activities | 368 | 39% | 23% | 41% | 36% | 814 | 11% | 19% | 46% | 34% |
| extra_bed | 450 | 48% | 22% | 43% | 35% | 1396 | 19% | 21% | 49% | 30% |
| out_of_bed | 153 | 16% | 14% | 30% | 56% | 281 | 4% | 13% | 28% | 59% |
| confusion | 51 | 5% | 47% | 33% | 20% | 72 | 1% | 39% | 43% | 18% |
| runny_nose | 339 | 36% | 53% | 35% | 12% | 2369 | 33% | 45% | 41% | 14% |
| blocked_nose | 345 | 37% | 41% | 44% | 14% | 1602 | 22% | 40% | 46% | 14% |
| sinus_pain | 240 | 25% | 30% | 48% | 21% | 912 | 13% | 33% | 48% | 19% |
| dry_cough | 497 | 53% | 42% | 43% | 15% | 1997 | 27% | 43% | 45% | 12% |
| white_phlegm | 171 | 18% | 47% | 40% | 12% | 729 | 10% | 49% | 41% | 10% |
| green_phlegm | 77 | 8% | 38% | 47% | 16% | 448 | 6% | 38% | 47% | 15% |
| loss_of_smell | 349 | 37% | 17% | 38% | 46% | 370 | 5% | 34% | 45% | 21% |
| loss_of_taste | 350 | 37% | 22% | 36% | 42% | 424 | 6% | 38% | 45% | 17% |
| sneezing | 352 | 37% | 64% | 26% | 10% | 1916 | 26% | 57% | 33% | 10% |
| sore_throat | 382 | 40% | 45% | 40% | 15% | 2574 | 35% | 43% | 43% | 14% |
| swollen_tonsils | 43 | 5% | 37% | 37% | 26% | 261 | 4% | 30% | 48% | 22% |
| swollen_glands | 81 | 9% | 43% | 46% | 11% | 459 | 6% | 45% | 40% | 15% |
| ear_pain | 125 | 13% | 45% | 43% | 12% | 643 | 9% | 47% | 42% | 12% |
| ear_fluid | 8 | 1% | 63% | 38% | 0% | 37 | 1% | 51% | 38% | 11% |
| sob | 250 | 26% | 34% | 48% | 18% | 673 | 9% | 35% | 46% | 18% |
| wheezing | 109 | 12% | 54% | 36% | 10% | 532 | 7% | 44% | 41% | 14% |
| chest_pain_nc | 107 | 11% | 43% | 45% | 12% | 251 | 3% | 45% | 41% | 14% |
| chest_pain_br | 97 | 10% | 39% | 41% | 20% | 226 | 3% | 45% | 41% | 14% |
| diarrhoea | 271 | 29% | 47% | 35% | 17% | 1048 | 14% | 43% | 37% | 20% |
| vomiting | 41 | 4% | 32% | 46% | 22% | 202 | 3% | 31% | 41% | 28% |
| nausea | 174 | 18% | 40% | 40% | 20% | 645 | 9% | 37% | 43% | 20% |
| abdo_pain | 118 | 13% | 42% | 42% | 16% | 699 | 10% | 31% | 45% | 24% |
| redeye | 65 | 7% | 46% | 49% | 5% | 317 | 4% | 49% | 40% | 10% |
| sticky_eye | 44 | 5% | 55% | 45% | 0% | 277 | 4% | 56% | 35% | 9% |
| eye_pain | 94 | 10% | 36% | 48% | 16% | 242 | 3% | 52% | 35% | 12% |
| eye_deter | 31 | 3% | 42% | 45% | 13% | 135 | 2% | 50% | 40% | 10% |
| rash_allover | 6 | 1% | 33% | 33% | 33% | 37 | 1% | 27% | 43% | 30% |
| rash_local | 44 | 5% | 43% | 50% | 7% | 194 | 3% | 42% | 41% | 17% |

Table S4. Duration of illness among swab-confirmed COVID-19 positive and negative illnesses, overall and stratified by age group and sex.

| Illness Type | Strata | | N | Mean | Standard Deviation | Range | | Percentile | | | |
|---|---|---|---|---|---|---|---|---|---|---|---|
| | | | | | | Lower | Upper | 25th | 50th | 75th | 90th |
| COVID+ve | Overall | Overall | 944 | 10.9 | 9.7 | 1 | 70 | 5 | 8 | 14 | 21 |
| | By age group | 0-15 | 76 | 6.6 | 5.4 | 1 | 25 | 2 | 6 | 9 | 14 |
| | | 16-24 | 54 | 7.4 | 7.9 | 1 | 56 | 3 | 6 | 9 | 13 |
| | | 25-44 | 270 | 9.7 | 8.4 | 1 | 63 | 4 | 7 | 12 | 18 |
| | | 45-64 | 379 | 12.5 | 10.5 | 1 | 70 | 6 | 9 | 15 | 25 |
| | | 65+ | 163 | 12.5 | 10.9 | 1 | 63 | 6 | 10 | 14 | 23 |
| | By sex | Male | 359 | 10.7 | 8.7 | 1 | 62 | 5 | 8 | 14 | 21 |
| | | Female | 517 | 11.6 | 10.6 | 1 | 70 | 5 | 9 | 14 | 23 |
| COVID-ve | Overall | Overall | 7269 | 5.9 | 7.3 | 1 | 182 | 2 | 4 | 7 | 12 |
| | By age group | 0-15 | 1038 | 5.1 | 4.4 | 1 | 36 | 2 | 4 | 7 | 11 |
| | | 16-24 | 246 | 5.1 | 5.5 | 1 | 45 | 2 | 4 | 7 | 10 |
| | | 25-44 | 1883 | 5.7 | 6.1 | 1 | 66 | 2 | 4 | 7 | 11 |
| | | 45-64 | 2779 | 6.0 | 8.0 | 1 | 182 | 2 | 4 | 7 | 12 |
| | | 65+ | 1262 | 7.1 | 9.2 | 1 | 166 | 2 | 5 | 7 | 14 |
| | By sex | Male | 2245 | 5.9 | 8.2 | 1 | 166 | 2 | 4 | 7 | 12 |
| | | Female | 4680 | 6.0 | 7.0 | 1 | 182 | 2 | 4 | 7 | 12 |

Figure S4. Distribution of illness duration* by age group among COVID-19 positive illnesses

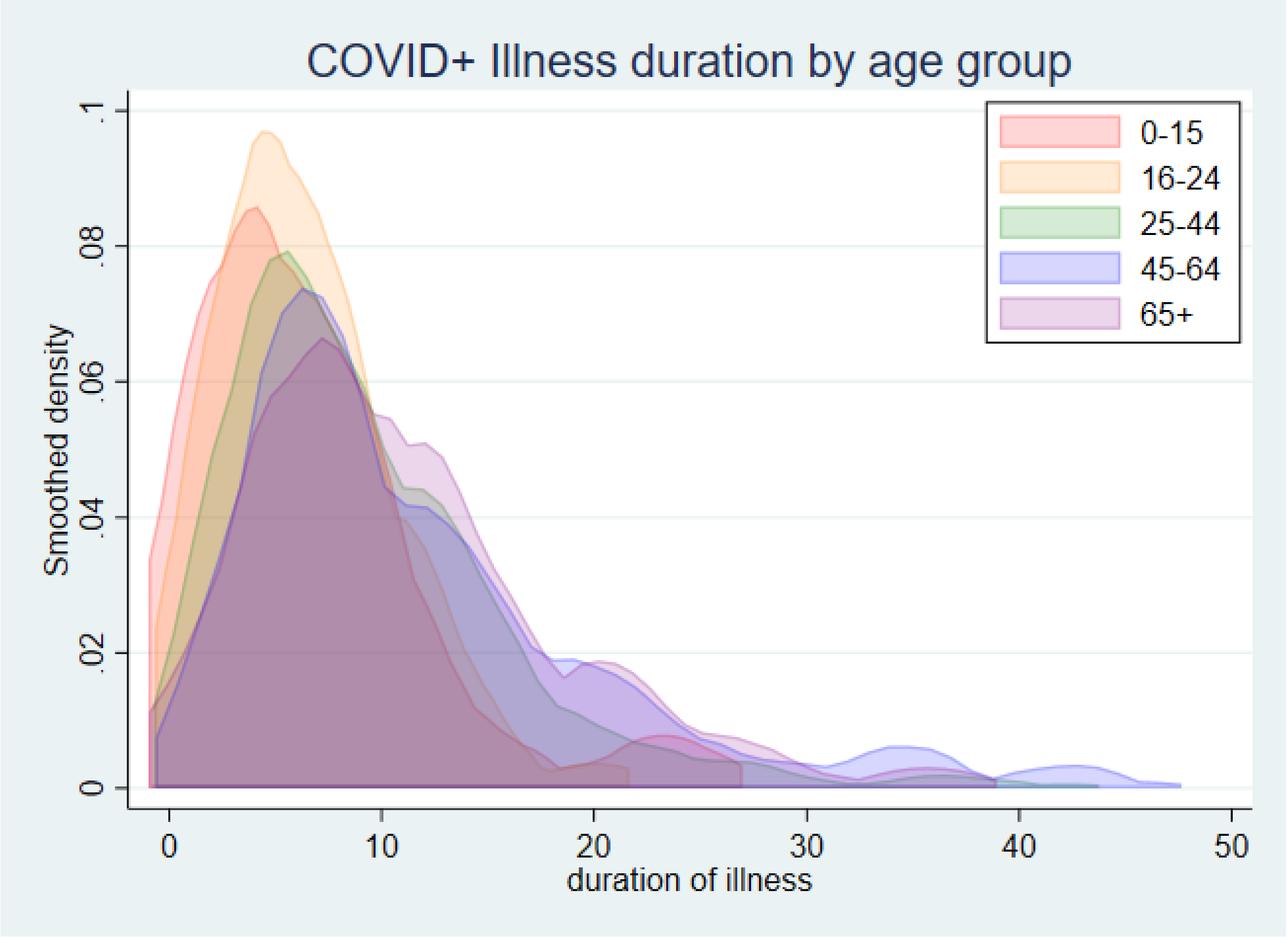


*Data limited to illnesses with duration <50 days.
